# Time to death and its determinants among neonates admitted to the Neonatal Intensive Care Unit (NICU) of Woldia Comprehensive Specialized Hospital, Amhara region, northeast Ethiopia: A five-year retrospective study

**DOI:** 10.1101/2024.03.10.24304055

**Authors:** Nuhamin Fentaw, Asmamaw Demis

## Abstract

**Background:** Despite an effort to reduce neonatal mortality, Ethiopia is still the leading among the countries with the highest neonatal death. While there have been studies conducted on the overall neonatal mortality rate, there is limited research specifically focusing on the time to death of neonates admitted to the Neonatal Intensive Care Unit (NICU).

**Objectives:** To assess the survival time of neonatal death (time to death) and its determinants for neonates admitted to the NICU of Woldia Comprehensive Specialized Hospital (WCSH).

**Methods:** A retrospective cohort study was conducted among 604 neonates admitted to the NICU of Woldia comprehensive specialized hospital from January 2018 to December 2022. The data were entered using Epi-Data version 4.6 and analysis was made using STATA version 14 software. To estimate failure probability, the Kaplan-Meier curve and life table estimate were calculated. The log-rank test was used to examine differences in death rates among different categories. The Cox proportional hazards model was employed to identify determinant variables. In the multivariate Cox analysis, variables with a hazard ratio p-value < 0.05 were considered statistically significant at a 95% confidence interval.

**Results:** The findings of the study indicate that a total of 604 neonates were included and observed for a cumulative period of 3416 neonate-days. The median time to death among the neonates was 14 days. Out of the reviewed cases, 117 (19.37%) neonates died during the study period. Have no ANC checkup (AHR: 1.68; 95% CI: 1.12-2.52), having bad obstetrics history (AHR: 1.99, 95% CI: 1.28-3.10), having sepsis (AHR: 1.87, 95% CI: 1.23-2.86) and having asphyxia (AHR: 1.64, 95% CI: 1.05-2.58) were increased the hazard of neonatal death.

**Conclusion:** The overall neonatal mortality incidence was high. ANC checkup, bad obstetrics history, and specific diagnoses, were found to significantly influence the hazard of neonatal mortality. Increase awareness and education programs for the community regarding the importance of ANC visits. Implement protocols for early identification and management of respiratory distress, sepsis and prenatal asphyxia is critical.

## Introduction

Neonatal period from birth to the first 28 days of life is the very critical to death (1). The probability of neonatal dying within this period of life is neonatal mortality (2). The death may be sub-grouped into early neonatal deaths, occurring during the first seven days of life (0-6 days) and late neonatal deaths, occurring after the seventh day but before the 28^th^ day of life (7-27 days) (3). The chance of dying is highest at this pivotal period of neonatal life (4).

Within a yearly, 2.6 million infants die before reaching one month old at the globally level (5). The continued burden of child mortality accounts an enormous loss of life alone in 2020, 5.0 (4.8-5.5) million children died before celebrating their fifth birthday. Half of those deaths, 2.4 (2.2-2.6) million were occurred among newborns. According to the current estimate of UN-IGME 47% of all under-five deaths occurred during the neonatal period (at the first 28 days of life) (6).

Greater number of these neonatal deaths appeared in low and middle income countries (7). Approximately 80% on neonatal mortality rate were concentrated in Sub-Saharan Africa and South Asia (8). Sub-Saharan Africa has the highest neonatal mortality rate in the world, at 27 (25–32) deaths registered as per 1000 live births in 2017 (9). As comparison, a child born from this region has a probability of eleven times more likely to die in the first month of life than a child born in a high-income countries (6). Similar studies have also shown that the risk of neonatal mortality is beyond 30 times higher in Sub-Saharan Africa than in the lowest mortality country (10). Neonatal mortality rates also significantly vary among the countries and regions with a huge toll of deaths attributed to the low-income countries (11). For instance, Ethiopia is among the countries with the highest neonatal mortality rate in Sub-Saharan Africa.

There are variances of causes in neonatal deaths in different regions and countries of economic development (12). Previous studies attempted to differentiate the likelihoods of neonatal mortality may be attributed to newborn, mother or health system factors (13–15). Factors associated with neonatal mortality includes birth weight, sex and neonatal infections such as neonatal sepsis, meningitis, respiratory distress syndrome(16), intraventricular hemorrhage, and persistent pulmonary hypertension (14) increased the risk of death. Maternal age, parity, birth interval, education; inadequate maternal knowledge on neonatal danger signs, complications at the time of delivery, history of abortion and home deliveries were associated causes which aggravate the hazard of death (7).

Between 1990 and 2020 the global neonatal mortality rate declined more slowly than mortality among children aged 1–59 months (1,6,8). There is a huge disparity of survival over regions and countries. Neonatal survival has significantly lower, for neonates born in Sub-Saharan Africa. The region continued to exposed the steepest odds of survival in the world (6).

Ethiopia is among these countries with lowest rate of neonatal survival records which placed at the 2^nd^ in the Sub-Saharan Africa next to Nigeria and the leading in east Africa (17). Despite different initiatives and implementations have been made to reduce neonatal death, still the mortality rate is high and not reduced as expected in Ethiopia. According to EDHS 2016 repot, neonatal mortality rate reported as 30.5 at 1^st^ day, 61.7 at 1^st^ week and 22.7 at 2^nd^ week of life. Within this the very slow neonatal mortality reduction progress, the sustainable development goal (SDG) would not be attained (18). In order to achieved the target, assessing the survival time of neonatal death, (time to death) and its determinants at the local level is very crucial and timely issue (19).

Many studies were conducted in different countries on the survival status and determinant of neonatal mortality but their main findings were emphasized on premature neonates. Likewise, studies done in Ethiopia are few and had focused on rate of neonatal mortality and little has been done on the time to death and its determinants. The methodology also they were used were cross sectional and few studies are retrospective with small number of observations and short period time interval (not more than 3 years). This could lack generalizability of the result and inability to determine the true mortality burden within the NICU.

Different studies revealed that the median time to death was 10 days (95%CI: 5, 19) with an overall incidence rate of death 6.81 per 1000 person-days observations (20). The cumulative proportion of surviving at the end of the 1^st^, 7^th^, 14^th^ and 21^st^ day were 96.1%, 75%, 69.9%, and 66.2% respectively (21). A retrospective study conducted in Addis Ababa among neonates found that the overall probability of survival of neonate was about 0. 612 for the follow-up days (22). Furthermore, a study conducted (23) showed, at the end of the follow-up, the cumulative probability of failure was 34.69 per 1000.

In sum, those studies showed inconsistent results and demonstrate the time to death of neonates admitted to NICU varies across different setting. Therefore, conducting a study on time to death and its determinants at the local level is timely issue to address the challenges of neonatal survival. Therefore, the main aim of this study was intended to determine the survival time of neonatal death (time to death) and its determinants among neonates admitted to the NICU of Woldia comprehensive specialized hospital.

## Methods

### Study setting

The study was conducted at Woldia Comprehensive Specialized Hospital (WCSH) which is located in Woldia town Amhara Region, Northeastern Ethiopia. It was established in 1961 and geographically located about 521km from Addis Ababa which is the capital city of Ethiopia and 360 km away from Bahir Dar, the capital city of Amhara.

The hospital serves more than 2.5 million people in Woldia town and neighboring regions and zones. It has different types of units: such as a medical ward, maternity ward, surgical ward, pediatric ward, ophthalmic ward and outpatient department, Intensive Care Unit (ICU), and other diagnostic services deliveries (24).

The Neonatal Intensive Care Unit (NICU) facility consists of 12 beds and is equipped with one X-ray machine, 12 radiant warmers, three rooms dedicated to term, preterm, and sepsis cases, as well as a separate room for kangaroo-mother care. Additionally, there are three rooms dedicated to maternity care. The NICU is staffed by a team comprising 10 nurses, two specialists, and two general doctors who provide comprehensive diagnosis and treatment for the admitted neonates.

On an annual basis, approximately 750 to 800 neonates were admitted to the NICU, seeking care for various health problems. The majority of admissions, accounting for 90% of cases were neonates referred from the labor ward, while the remaining 10% were referred from other birth centers and health facilities.

### Study design and period

A retrospective cohort study was conducted on neonates who were admitted to the Neonatal Intensive Care Unit (NICU) at Woldia Comprehensive Specialized Hospital (WCSH) between January 2018 to December 2022.

### Source population

The source population of this study consisted of all neonates who were admitted to the Neonatal Intensive Care Unit (NICU) at Woldia Comprehensive Specialized Hospital (WCSH).

### Study population

The study included all neonates who were admitted to the Neonatal Intensive Care Units (NICU) at Woldia Comprehensive Specialized Hospital (WCSH) between January 1st, 2018 and December 30th, 2022, and whose records were accessible in the admission and discharge registry.

### Inclusion criteria

Only babies aged 0-28 days will be included in the survey. All medical records of neonates who were hospitalized in Neonatal Intensive Care Units (NICU) from January 2018 to December 2022 in Woldia Comprehensive Specialized Hospital (WCSH) will be included in the study.

### Exclusion criteria

Neonates with lost or incomplete registered NICU logbooks or medical charts (records like admission data, discharge date, missed diagnosis), and neonates whose discharge summary sheet did not clearly show whether they were alive or died were excluded from this study at the time of data reviewing.

### Sample size determination and sampling techniques

The sample size was determined using STATA Version 14 statistical software assuming: confidence interval 95%, power 80%, the hazard ratio 3.52 not crying at birth, 0.03469 probability of neonatal death, 0.5 variability and 5% contingency from previous studies conducted in Ethiopia (23). The final sample size yields by considering not crying at birth was 604. Sampling frame was prepared from the medical charts of the neonates that were hospitalized from January 2018 to December 2022 in the NICU. The sample size was proportionally allocated to each year for optimal allocation. Finally, the study subjects were chosen using straightforward random selection techniques and a lottery system.

### Data collection methods and tools

To facilitate data collection, structured questionnaire checklists were developed by reviewing previous studies (25–28). Prior to data collection, the questionnaires’ contents (cheek list) underwent validation, and a pre-test was conducted to ensure their effectiveness in similar scenarios. Subsequently, improvements and adjustments were made accordingly.

The medical records of eligible neonates were manually searched using their unique medical record numbers. Information pertaining to the neonates was extracted from their medical records, Furthermore, the medical records of the mothers also traced to obtain maternal factors such as parity, gravidity, antenatal care (ANC) follow-up, age, and medical diagnoses during pregnancy, which were reviewed from delivery.

### Study variables

Dependent variable: The dependent variable of this study was time to event (time to death). Independent variables: Different independent variables or explanatory variables were; Socio-demographic factors: maternal age, residence, and sex and age of neonate. Maternal obstetrics factors: ANC follow-up, complication during pregnancy and birth, parity, gravidity, maternal Rh factor, onset of labor, history of bad obstetrics. Neonatal factors: neonatal sex, age, gestational age, birth weight, Apgar score, cried at birth. Neonatal medical condition: neonatal anemia, asphyxia, neonate jaundice, neonatal sepsis, RDS (respiratory distress), neonatal hypoglycemia, necrotizing enterocolitis, meningitis, MAS (Meconium aspiration) and TTN (transient tachypnea of the newborn).

### Operational Definition

**Neonatal mortality (NM):** is defined as the probability of dying within the first month of life and expressed as neonatal deaths per 1000 live births (29). Neonatal deaths were categorized as:

**Early neonatal mortality (ENM):** Death of a live-born infant during the first seven completed days (168 hours) of life.

**Late neonatal mortality (LNM):** Death of a live-born infant after 7 completed days but before 28 completed days of life (30).

**Censored:** didn’t know survival time exactly due to study ends, loss to follow-up, withdrawal or being survived from study.

**Event:** neonatal death after NICU admission.

**Time to death:** it is the time from admission at NICU to the occurrence of the outcome/event (discharge/death) (31). It is the time (date) difference between admission and time of event occurred.

**Time scale:** days from admission of a neonate to the occurrence of an outcome.

**Time origin:** date of admission or start time of the cohort.

**Congenital anomaly:** is a structural and functional abnormality that presents at birth. It is body deformity from the birth believed to have impact on health of the baby (32).

**Sepsis:** Record of infection or sepsis diagnosed either clinically or with culture by professionals during admission of the neonate and as possible causes of death and designated on chart (33).

**Apgar score:** is a simple evaluation system including five easily identifiable components such as heart rate, respiratory effort, muscle tone, reflex/activity, and color (10).

**Low birth weight (LBW):** is defined as a birth weight of a live born infant of less than 2500 g, very low birth weight (VLBW) is less than 1500 g, and extremely low birth weight (ELBW) is less than 1000 g (34).

**Prematurity:** is defined by a gestational age less than 37 weeks. Based on gestational weeks, prematurity can be sub-grouped as; ‘extremely preterm’ (less than 28 weeks), ‘very preterm’ (28 to 31 weeks), and ‘moderate to late preterm’ (32 to 36 weeks) (35).

### Data quality control and analysis

Prior to commencing the actual data collection, a preliminary review was conducted on a 5% of the study subject with the study period to validate the data extraction tools. Training was provided to both data collectors and supervisors involved in the data collection and extraction process. Furthermore, a thorough check was conducted to ensure the completeness, consistency, clarity, and accuracy of the data before entering it into the software.

The collected data underwent a thorough examination to ensure completeness, consistency, and accuracy. The data were entered into Epi-Data Manager version 4.6 and exported to STATA version 14 software for analysis. Percentages, frequencies and medians were utilized to describe the sociodemographic variables. To estimate the probability of neonatal death during the follow-up period, the Kaplan-Meier curve and life table estimate were computed. Furthermore, the log-rank test was employed to identify any statistical differences between groups.

Cox proportional hazard model was used for survival analysis to observe the association between various independent variables and time to death. Univariate analysis was initially employed to identify variables associated with the survival time of neonatal death. Variables with a p-value less than 0.25 were considered statistically significant and included in the multivariable Cox proportional hazard regression model as significant determinants of time to death. Additionally, the overall model fitness was assessed using the Schoenfeld residual global and graphical tests. The level of statistical significance was set at p < 0.05 to determine the significance of the findings.

### Ethical consideration

Ethical clearance was obtained specifically from the Institutional review board of Woldia University college of health science. Additionally, authorization to conduct the study and ethical clearance were obtained from the hospital authorities and the ethical committee. Throughout the data collection process, utmost care was taken to ensure the confidentiality and privacy of the patients’ medical records.

## Results

### Maternal sociodemographic and obstetrics characteristics

The majority, 415 (68.71%), resided in rural areas outside of Woldia town. In terms of parity, 269 (44.54%) were primiparous, while over 40% were multiparous women. Among the subjects, 335 (55.46%) were multigravida, and 269 (44.54%) were primigravida.

Out of the total observations, 475 (78.68%) mothers had received antenatal care (ANC) check-ups, and only 142 (23.51%) received the tetanus toxoid (TT) vaccine once throughout their pregnancy. The Rh-status of the majority of mothers, 446 (73.84%), was positive.

Maternal illnesses were reported by 69 (11.42%) during pregnancy, 185 (30.63%) experienced pregnancy complications, and 122 (20.20%) encountered labor complications. Among the neonates’ mothers, only 71 (11.75%) had a history of bad obstetrics (Table 1).

**Table 1.**
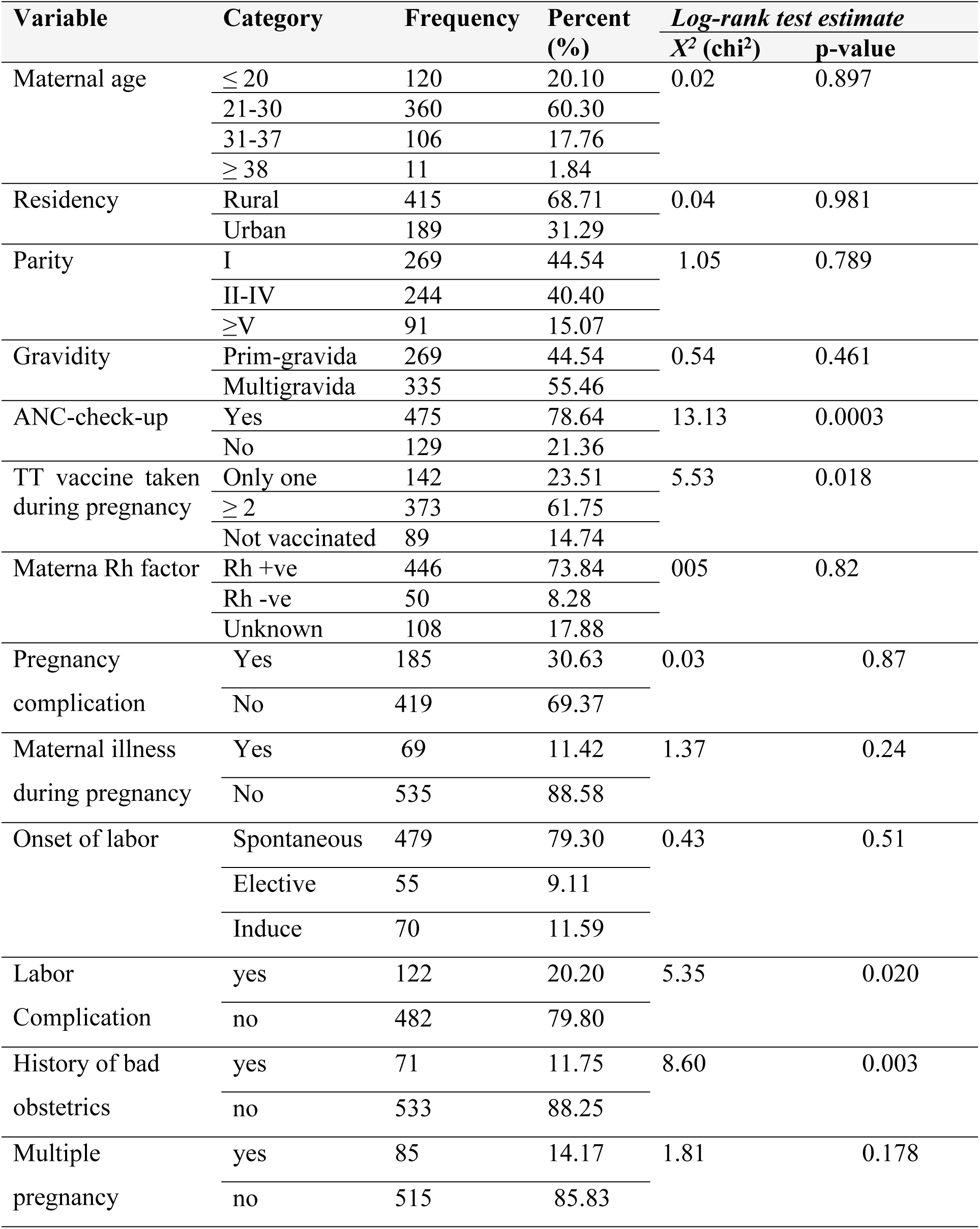
Materna demographic and obstetrics characteristics of neonates admitted to the NICU of Woldia comprehensive Specialized hospital from January 2018 to December 2022.

### Neonatal demographic characteristics and medical conditions

During the specified period from January 2018 to December 2022, a total of 3,136 neonates were admitted to the Neonatal Intensive Care Units (NICU) of Woldia comprehensive specialized hospital. Among them, 604 selected patient chart records were reviewed. Of the studied subject, 292 (48.34%) were male neonates. More than half, 487 (80.63%), were less than 7 days old. In terms of gestational age, 60 (9.93%) were extremely preterm (born before 32^+6^ weeks), 205 (33.94%) were preterm (born between 33 and 37 weeks), 22 (3.64%) were post-term, and the majority, 317 (52.69%), were delivered between 37 and 41^+6^ weeks of gestational age.

Analysis results indicated that 316 (52.32%) of the neonates had a normal birth weight between 2501 and 4000g. Extremely low birth weight (less than 1500g) and low birth weight (less than 2500g) accounted for 93 (15.40%) and 139 (23.01%) cases, respectively. Among the admitted neonates, the 1^st^ and 5^th^ minute APGAR scores matched between 4 and 6 for the majority, with 262 (43.38%) and 274 (40.89%) cases, respectively. APGAR scores below three were observed in 148 (24.50%) and 101 (16.72%) cases for the 1^st^ and 5^th^ minutes, respectively. A total of 318 (52.65%) neonates cried during birth, and 355 (58.77%) were exclusively breastfed, while only 43 (7.12%) initiated breastfeeding within 30 minutes after delivery (Table 2).

**Table 2.** Demographic characteristics and clinical condition of neonates admitted to the NICU of Woldia Comprehensive specialized Hospital from January 2018 to December 2022.

| Variable | Variable Category | Frequency | Percent (%) | Log rank test estimate |  |
| --- | --- | --- | --- | --- | --- |
|  |  |  |  | X<sup>2</sup> | p-value |
| Neonatal sex | Male | 292 | 48.34 | 5.14 | 0.023 |
|  | Female | 312 | 51.66 |  |  |
| Age | 0-6 days | 487 | 80.63 | 1.95 | 0.162 |
|  | 7-27 days | 117 | 19.37 |  |  |
| Gestational age | ≤ 32<sup>+6</sup> weeks | 60 | 9.93 | 9.66 | 0.001 |
|  | 33-36<sup>+6</sup> weeks | 205 | 33.94 |  |  |
|  | 37-41<sup>+6</sup> weeks | 317 | 52.48 |  |  |
|  | ≥ 42 weeks | 22 | 3.64 |  |  |
| Birth weight | ≤ 1500 g | 93 | 15.40 | 93.70 | 0.000 |
|  | 1501-2500 g | 139 | 23.01 |  |  |
|  | 2501-4000 g | 316 | 52.32 |  |  |
|  | ≥ 4000 g | 56 | 9.27 |  |  |
| 1<sup>st</sup> minute Apgar score | ≤ 3 | 148 | 24.50 | 13.11 | 0.0003 |
|  | 4-6 | 262 | 43.38 |  |  |
|  | ≥ 7 | 157 | 25.99 |  |  |
|  | Unknown | 37 | 6.13 |  |  |
| 5<sup>th</sup>-minute Apgar score | ≤ 3 | 101 | 16.72 | 8.96 | 0.002 |
|  | 4-6 | 247 | 40.89 |  |  |
|  | ≥ 7 | 219 | 36.26 |  |  |
|  | Unknown | 37 | 6.13 |  |  |
| Health facility visit after the neonate getting sick |  |  |  |  |  |
|  | Before 3Hrrs. | 357 | 59.11 | 4.24 | 0.039 |
|  | After 3Hrs. | 247 | 40.89 |  |  |
| Baby cried at birth | yes | 318 | 52.65 | 16.12 | 0.0001 |
|  | no | 286 | 47.35 |  |  |
| Body temperature | <36.5 | 266 | 44.04 | 4.49 | 0.03 |
|  | 36.5-37.5 | 281 | 46.52 |  |  |
|  | >37.5 | 57 | 9.44 |  |  |
| Breast-feeding initiation time | within-30 min | 43 | 7.12 | 5.63 | 0.040 |
|  | within-1 hr. | 142 | 23.51 |  |  |
|  | after 1 hr. | 419 | 69.37 |  |  |
| Exclusive breast-feeding | yes | 355 | 58.77 | 1.39 | 0.238 |
|  | no | 249 | 41.23 |  |  |

The common medical problems among the admitted neonates included neonatal anemia (58, 9.6%), prenatal asphyxia (217, 35.93%), neonatal sepsis (183, 30.30%), neonatal hypoglycemia (57, 9.44%), respiratory distress (87, 14.44%), meconium aspiration syndrome (134, 22.19%), transient tachypnea of the newborn (51, 8.44), birth trauma (81, 13.41%), necrotizing enterocolitis (37, 6.13%), and meningitis (36, 5.96%) (Table 3).

**Table 3.**
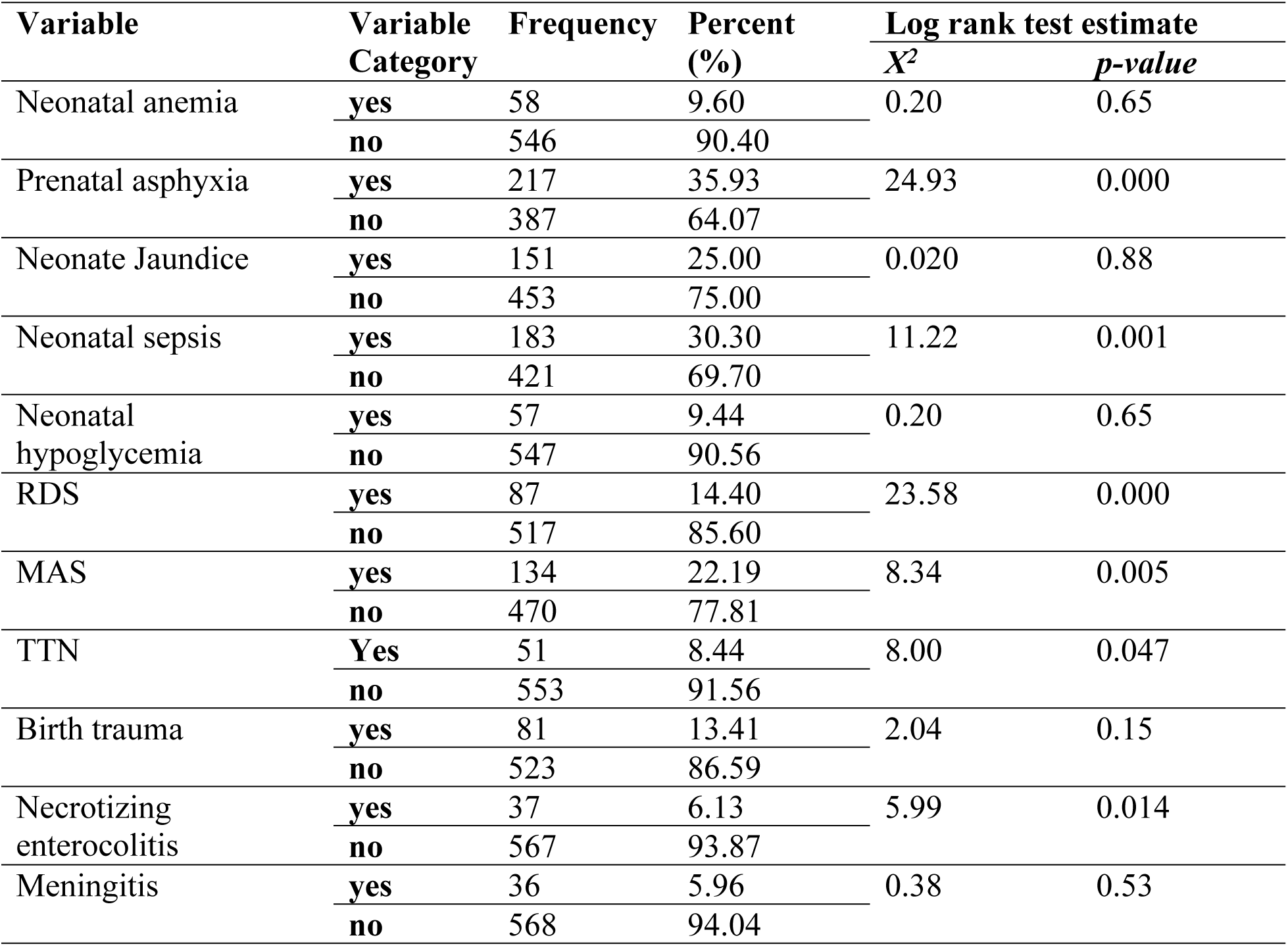
Medical diagnosis of neonates admitted to the NICU of Woldia comprehensive specialized hospital from January 2018 to December 2022.

### Follow up outcome and time to death

In this study, a total of 604 neonates admitted to the Neonatal Intensive Care Unit (NICU) of Woldia comprehensive specialized hospital (WCSH) from January 1, 2018, to December 30, 2022, were followed for a total of 3,416 neonatal days over a period from admission to discharge/death. The median follow up was 5 days with a minimum of 1 day and the maximum of 28 days observations. The overall incidence of neonatal mortality was 34.3 per 1000 neonates-days observations. This leads to 194 per 1000 live births of overall neonatal death rate.

Among the reviewed cases, 117 (19.37%) resulted in death, with 3.14% occurring despite medical treatment options and 2.81% requiring referral to other hospitals. The majority, more than 70%, were alive and discharged to home during the follow-up period (Fig 1).

**Fig 1.**
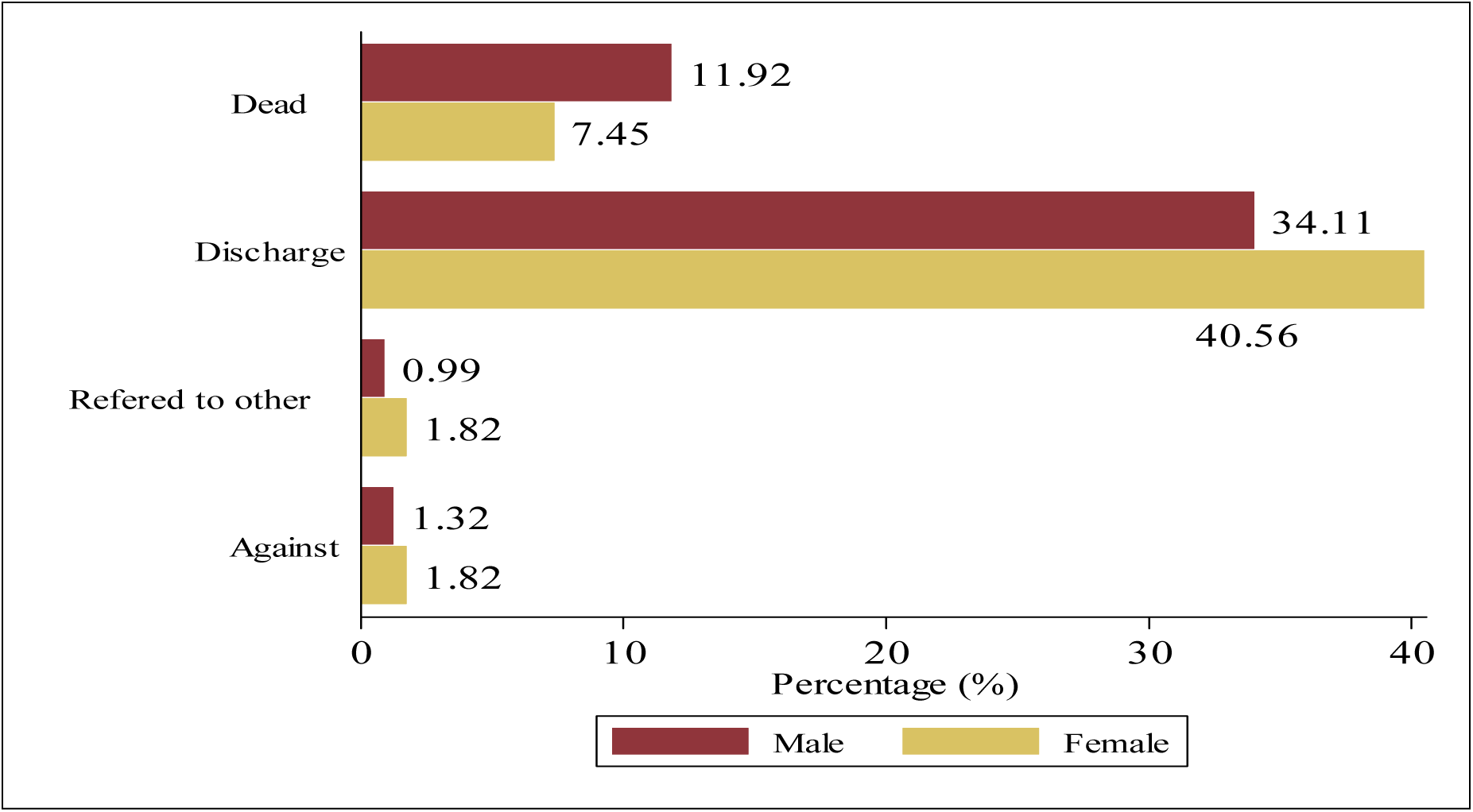
Outcomes of neonates admitted to the NICU of Woldia comprehensive specialized hospital from January 2018 to December

Out of the 117 deaths, the majority (63.24%) occurred within the first 7 days, 34.18% occurred between 7 and 14 days, and 2.56% occurred between 14 and 28 days of neonatal life. Based on the life table estimate, the cumulative probability of death at the end of each follow-up day (7^th^, 14^th^ and 28th days) was 0.17, 0.42 and 0.52, respectively (Table 4).

**Table 4.** Failure Probability (Life table estimates of the cumulative progression to death) among neonates admitted to the NICU at Woldia Comprehensive Specialized Hospital from January 2018 to December 2022.

| Time<br>(In day) | Neonates entering<br>interval | Failure event<br>(death) | Failure probability (95% CI) |
| --- | --- | --- | --- |
| 0-7 | 604 | 74 | 0.17(0.14-0.21) |
| 7-14 | 197 | 40 | 0.42(0.35-0.50) |
| 14-21 | 25 | 3 | 0.52(0.41-0.65) |
| 21-28 | 7 | 0 | 0.52(0.41-0.65) |

Furthermore, based on the Kaplan-Meier failure estimate curve, the probability of neonatal death during the first 7 consecutive follow-up days showed a steep rise, indicating a higher probability of neonatal death. Similarly, between 7 and 14 days, the curve suggests an initially high hazard of death. However, for the remaining follow-up period, the curve shows a slight rise and goes to flat, indicating a lower hazard of death (Fig 2).

**Fig 2.**
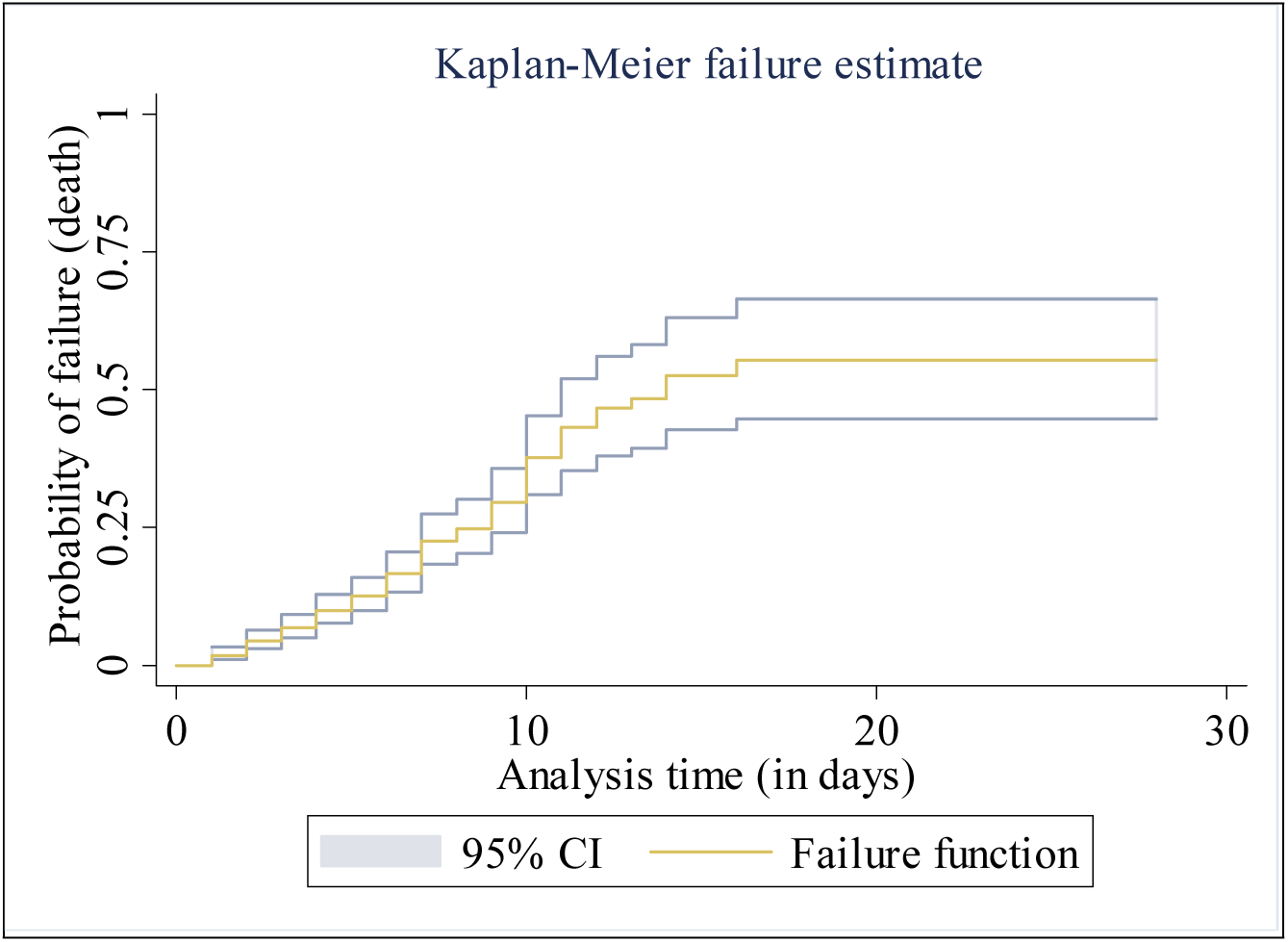
Overall Kaplan Meier failure estimate of neonates admitted in NICU at Woldia comprehensive specialized hospital from 1^st^ January 2018 to December 30^th^ 2022

### Comparison of failure function for different groups of neonates

Neonates born from mothers who experienced labor complication during birth have a greater probability of failure than those of birth who did not have complication. Neonates born from mothers who experienced labor complication during birth, has lower survival prognosis. For neonates born to moms who had a labor complication, the median time to death (10 days) was shorter than the median time to death (16 days) for neonates born to mothers who did not have a labor complication (Fig 3).

**Fig 3.**
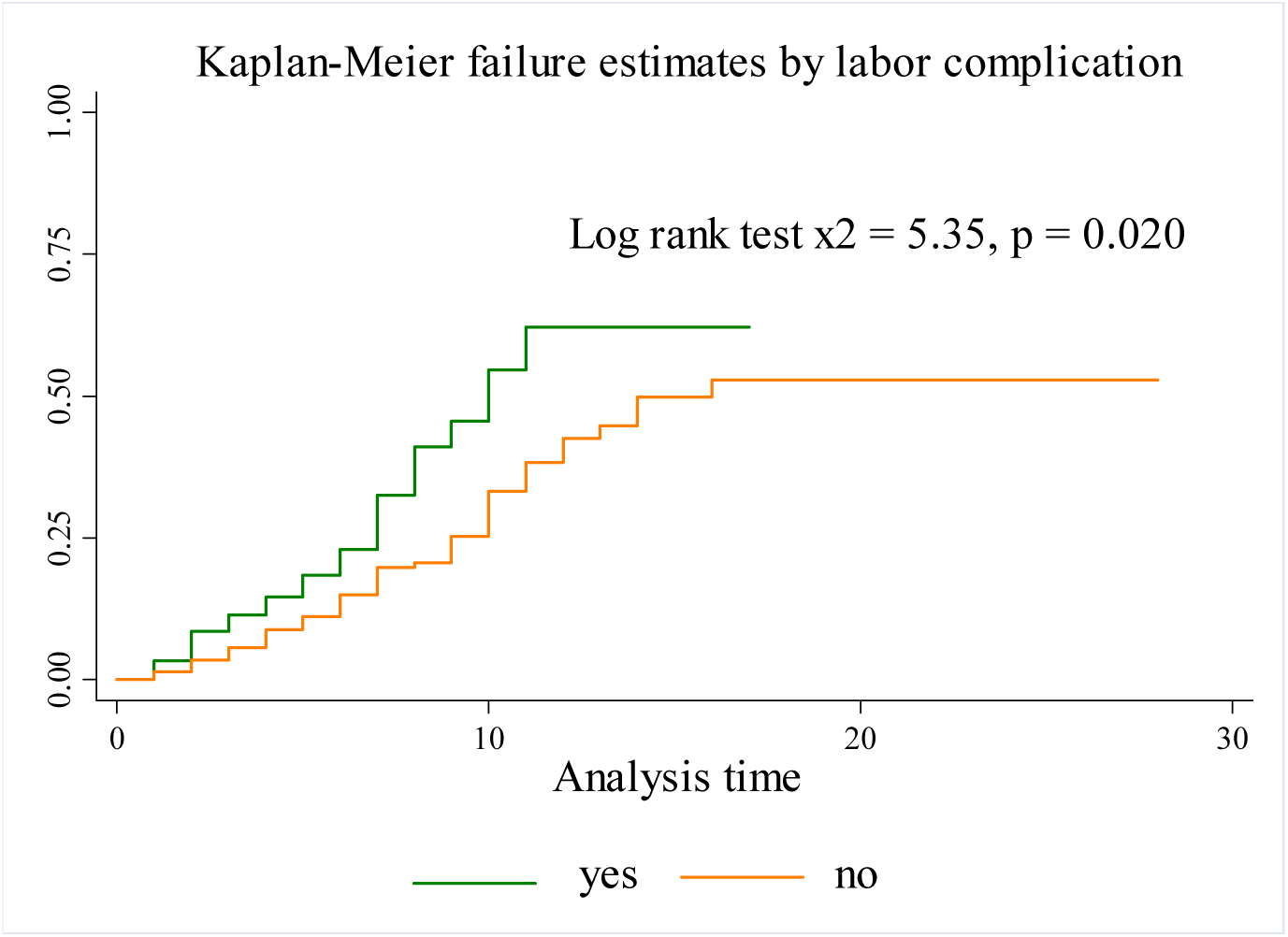
Kaplan Meier failure estimate comparison of time to death of neonates with groups of labor complication for neonates admitted to the NICU of Woldia Comprehensive specialized hospital from 1^st^ January 2018 to December 30^th^ 2022

This finding noticed that, the KM-failure curve for neonates born from mothers who did not take ANC recommendation consistently higher than the KM failure curve for whose mother has taken. Additionally, the median hazard time of death for neonates born from mothers who did not attend ANC during their indexed pregnancy was (days 10, 95% CI 6-16) compared with neonates born from mothers who received ANC service (Fig 4).

**Fig 4.**
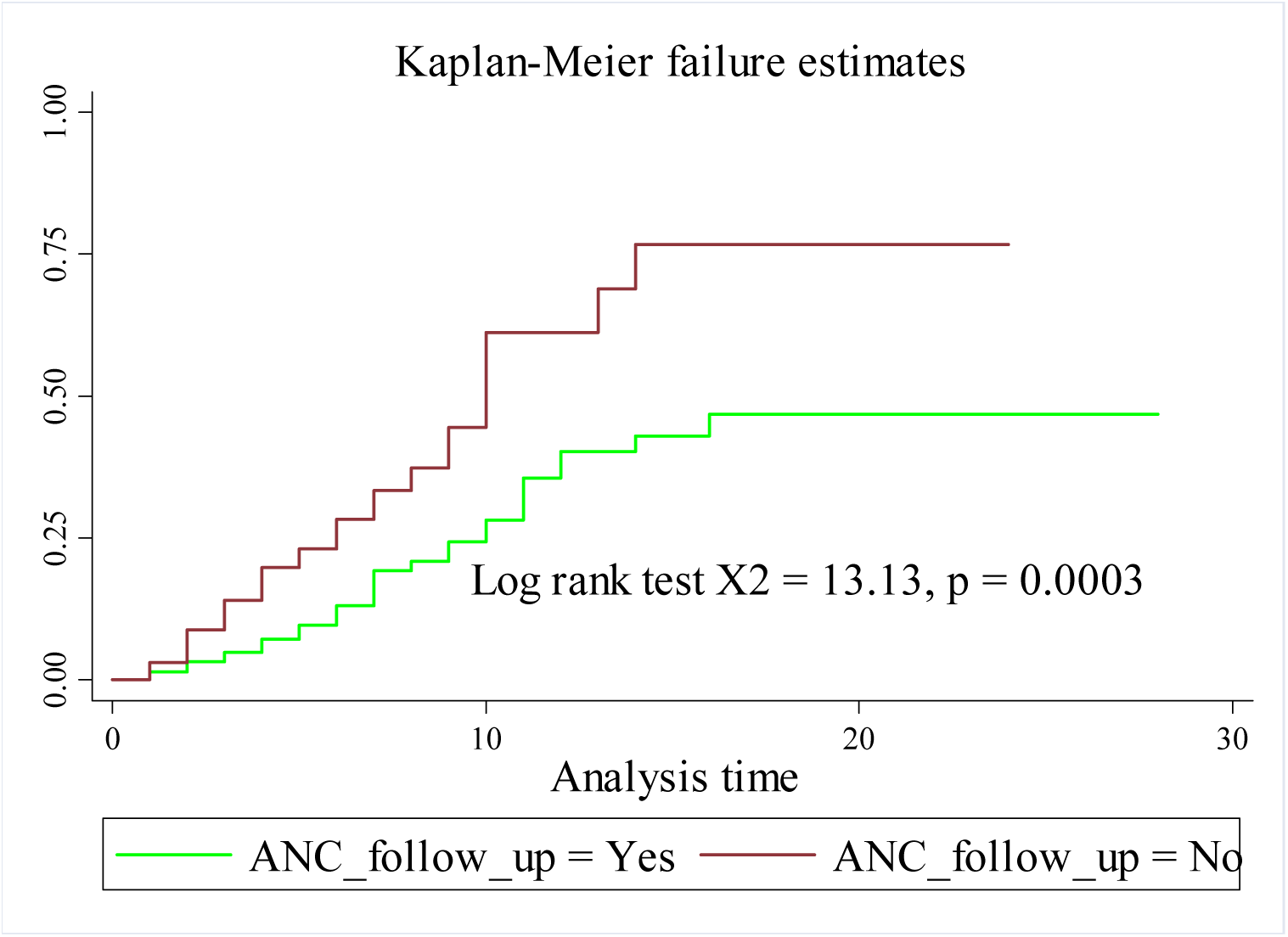
Kaplan Meier failure estimate comparison of time to death of neonates with groups of ANC-checkup for neonates admitted to the NICU of Woldia comprehensive specialized hospital from 1^st^ January 2018 to December 30^th^ 2022

This study revealed that failure probability of neonates born from among the groups of bad obstetrics history of the mothers was significantly different. Babies born from moms with bad obstetrics history, the median survival time was 11 days which was shorter than 16 days the time to survive neonates born to mothers who did not have bad obstetrics history (Fig 5).

**Fig 5.**
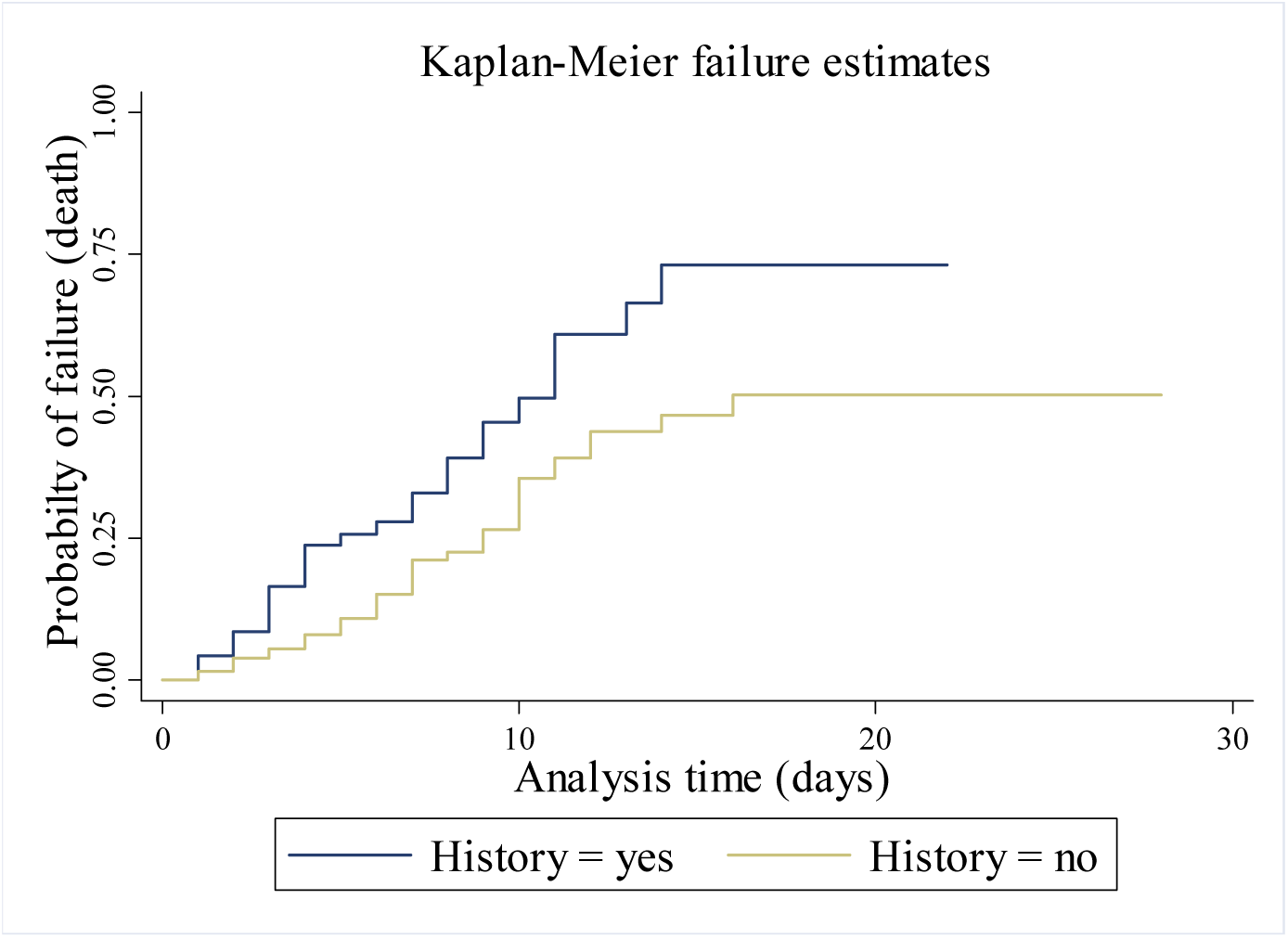
Comparing time to death of neonates with categories of maternal bad obstetrics history for neonates admitted at the NICU of Woldia comprehensive specialized hospital between 1st January 2018 and December 30th 2022

Likewise, according to the Kaplan Meier failure estimate, extremely low birth weight (less than 1500 g) and low birth weight (1501 to 2500 g) had higher hazard of death than those of normal birth weight neonates. This is possibly to say that the proportion of failure among neonates with extremely low and low weight at admission were higher relative to those of with normal weight. The hazard of death was significantly higher for those extremely low and low birth weight during the first week of life (early neonatal life) (Fig 6).

**Fig 6:**
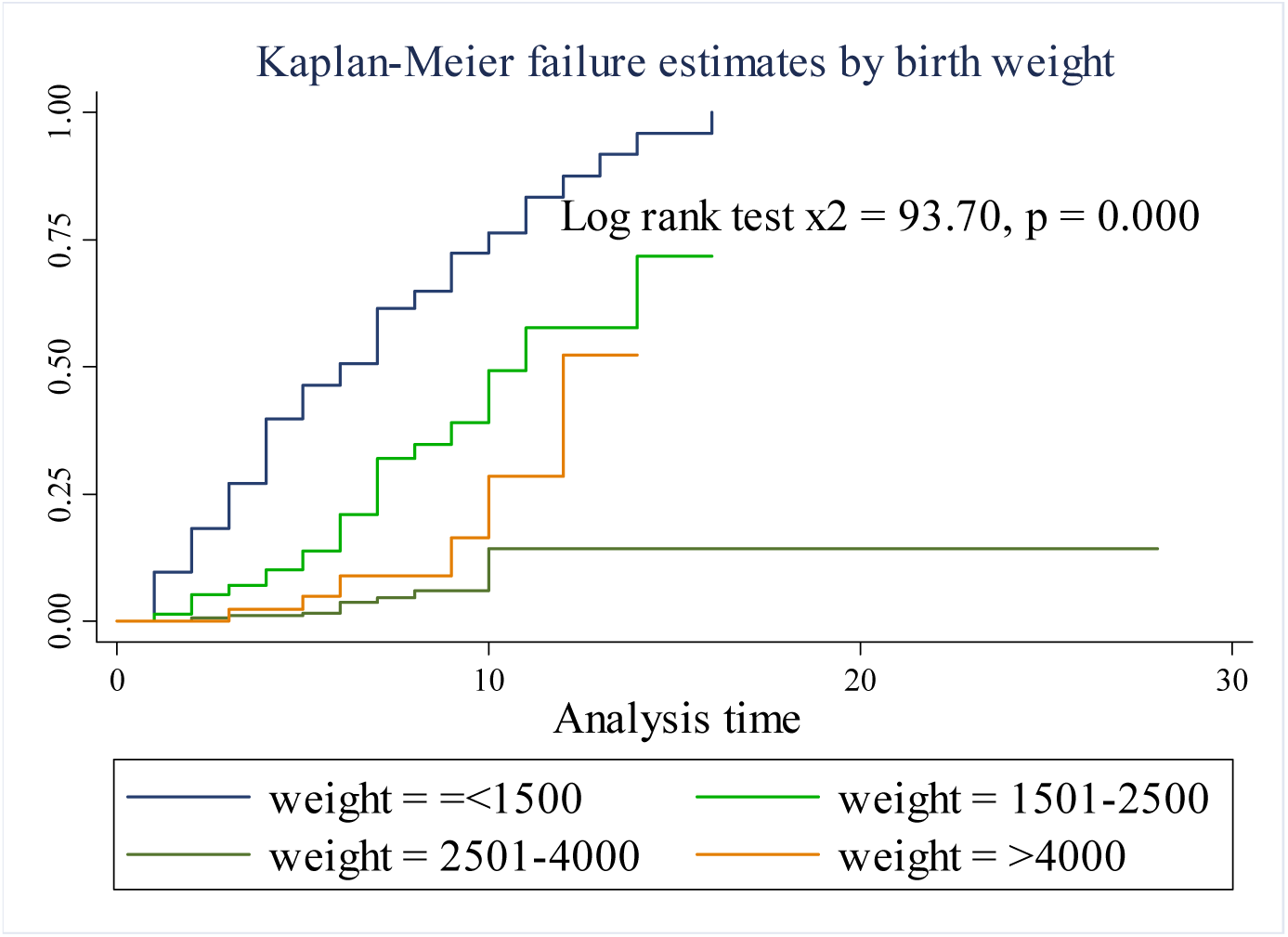
Kaplan Meier failure estimate comparison of time to death difference among the groups of birth wight for neonates admitted to the NICU of Woldia comprehensive specialized hospital from 1^st^ January 1018 to December 30^th^

Babies who were diagnosed and positive with asphyxia had a greater hazard of dying compared to babies who were not. The twenty-five-percentile hazard time of death for babies with asphyxia was 7 days which was shorter than the twenty-five-percentile hazard time of death for babies without asphyxia (10 days), with an incidence rate of death 55.6/1000 (Fig 7).

**Fig 7:**
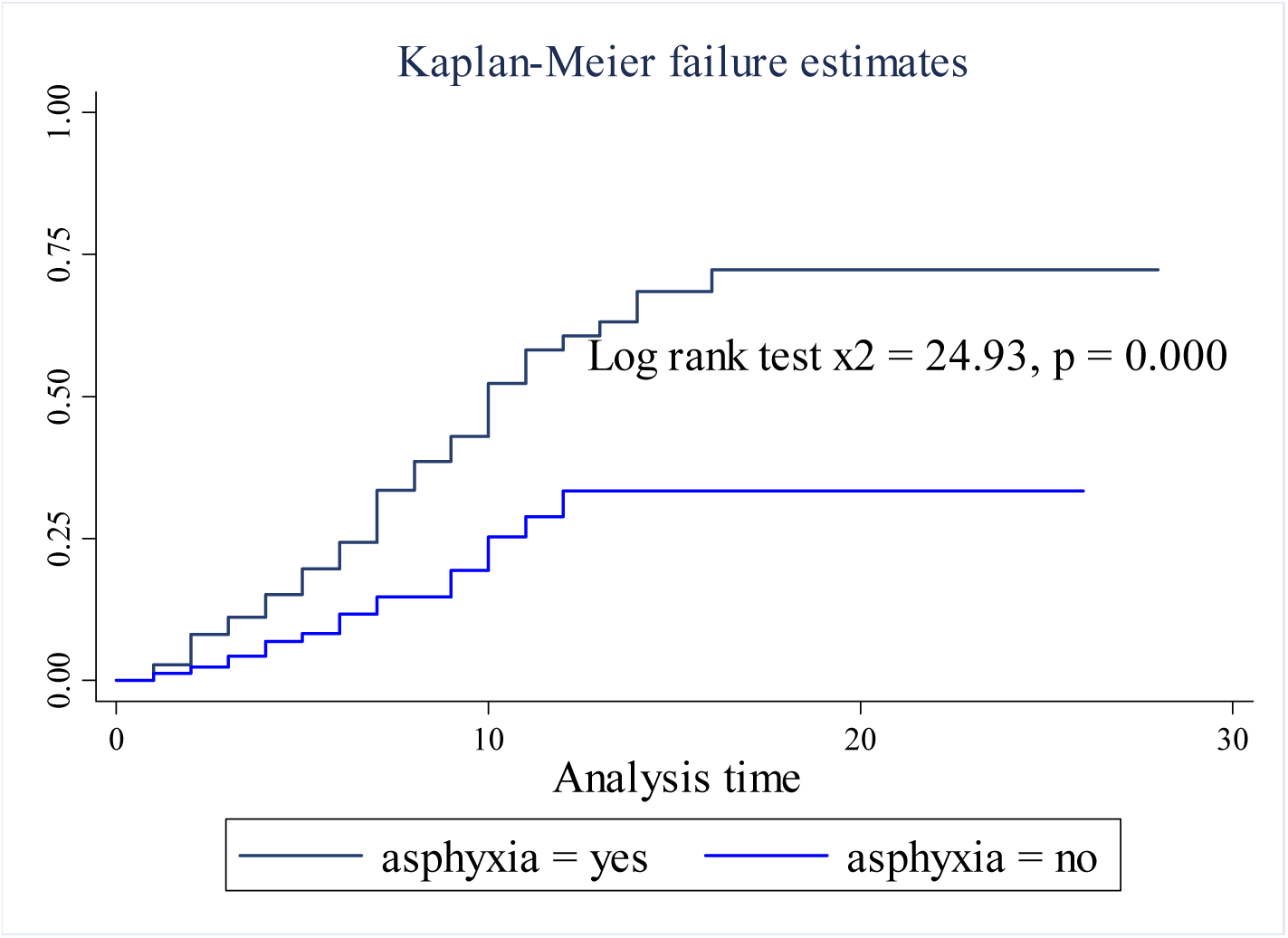
Kaplan Meier failure estimate comparison of time to death among the group of asphyxia for neonates admitted to the NICU of Woldia Comprehensive specialized hospital from 1^st^ January 2018 to December 30^th^ 2022

### Determinants of Mortality and Time to Death of Neonates Admitted to NICU

Both univariate and multivariable Cox proportional hazard models were employed to determine the factors influencing the time to death and mortality of neonates admitted to the Neonatal Intensive Care Unit (NICU) during the study period. The results of the univariate Cox regression analysis indicated that several independent variables were significantly associated with mortality and the time to death of neonates in the NICU.

These variables include ANC check-up, labor complications, maternal history of bad obstetrics, neonatal sex, gestational age, birth weight, health facility visits after neonatal illness occurred, body temperature, breastfeeding initiation time, crying at birth, and the presence of diseases such as prenatal asphyxia, neonatal sepsis, respiratory distress, and necrotizing enterocolitis.

The multivariable Cox proportional hazard model was used to further examine the associations while adjusting for other potential confounding factors (Table 5). However, in the final multivariable Cox proportional hazard analysis, after adjusting for potential confounders, only certain variables remained significant determinants of mortality and the time to death for neonates admitted to the NICU.

**Table 5.**
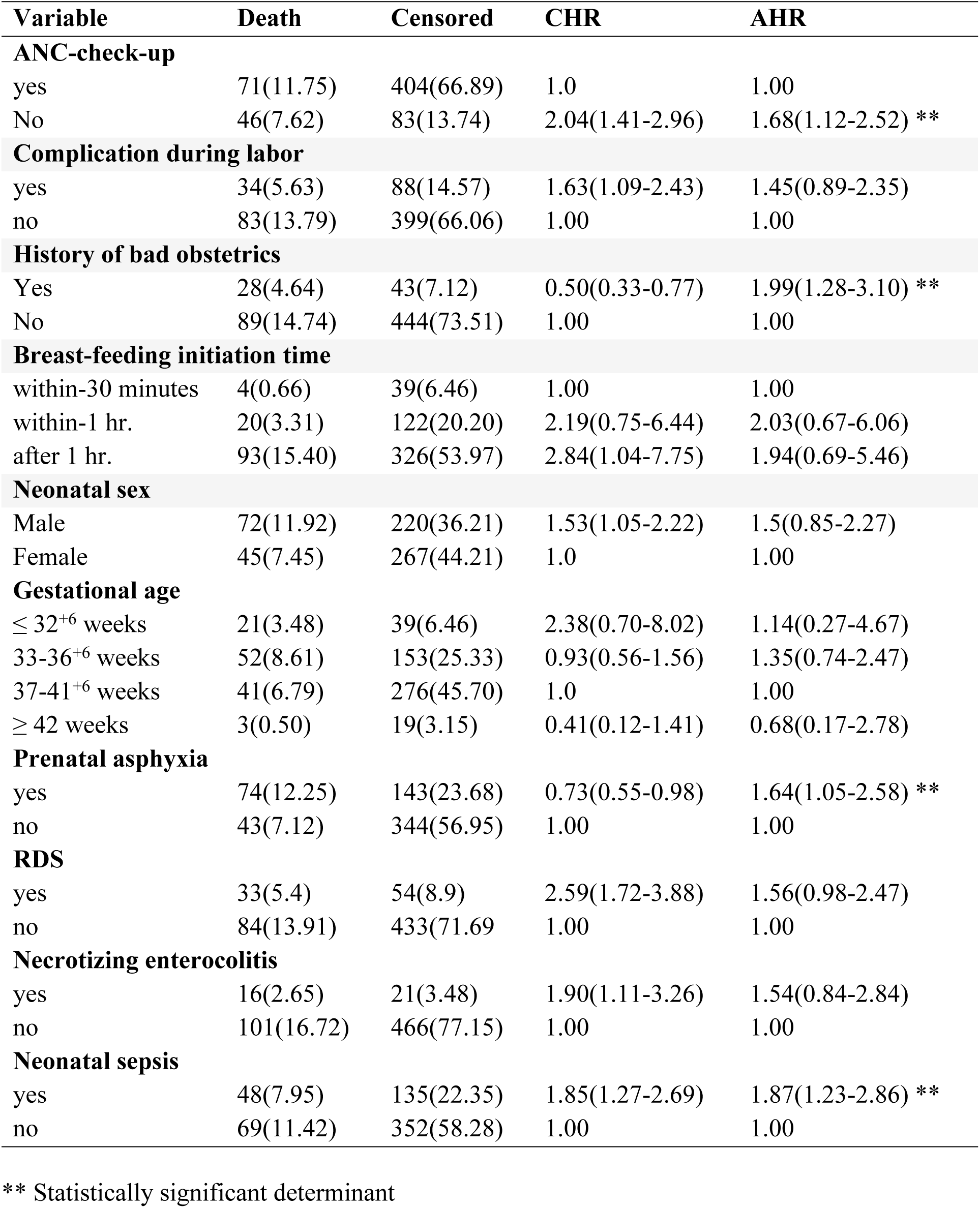
Univariate and multivariable Cox-proportional hazard for determinants of neonatal mortality neonates admitted to NICU at WCSH from January 2018 to December 2022.

Specifically, variables including ANC check-up, maternal history of bad obstetrics, neonatal sepsis, and prenatal asphyxia. These factors retained their significance even after controlling for confounding variables (Table 5).

Multivariable Cox proportional hazard regression analysis revealed that neonates born to mothers who did not attend ANC checkups during pregnancy were found to have a 1.7 times higher hazard of death compared to neonates born to mothers who received ANC checkups (AHR 1.68, 95% CI 1.12-2.52). Among the admitted neonates, those born to mothers with a history of bad obstetrics had an 83% higher hazard of death compared to neonates whose mothers had no history of bad obstetrics (AHR 1.99, 95% CI 1.28-3.10).

Similarly, neonates with neonatal sepsis had a 1.87 times higher hazard of mortality compared to non-septic neonates (AHR 1.87, 95% CI (1.23-2.86). Neonates with asphyxia were found to have 1.64 times higher hazard of death (AHR 1.64, 95% CI 1.05-2.58) (Table 5).

### Diagnosis of the Cox proportional hazard assumption

By using the Schoenfeld residuals test, the Cox proportional hazard assumption was statistically assessed for each covariate. The result of the assessment explained that the p-value for each variable and the whole covariates were above the permissible limit (p = 0.05). In addition to statistical assumption test, the model also graphically assessed.

The graph clearly demonstrated that for all models, plots of Cox-Snell residuals versus estimated cumulative hazard function were closer to the line across the origin. The graphic for the Weibull baseline distribution follows and draws straight lines with the origin. This demonstrated that it was suitable and appropriate and fitted for the dataset (Fig 8).

**Fig 8:**
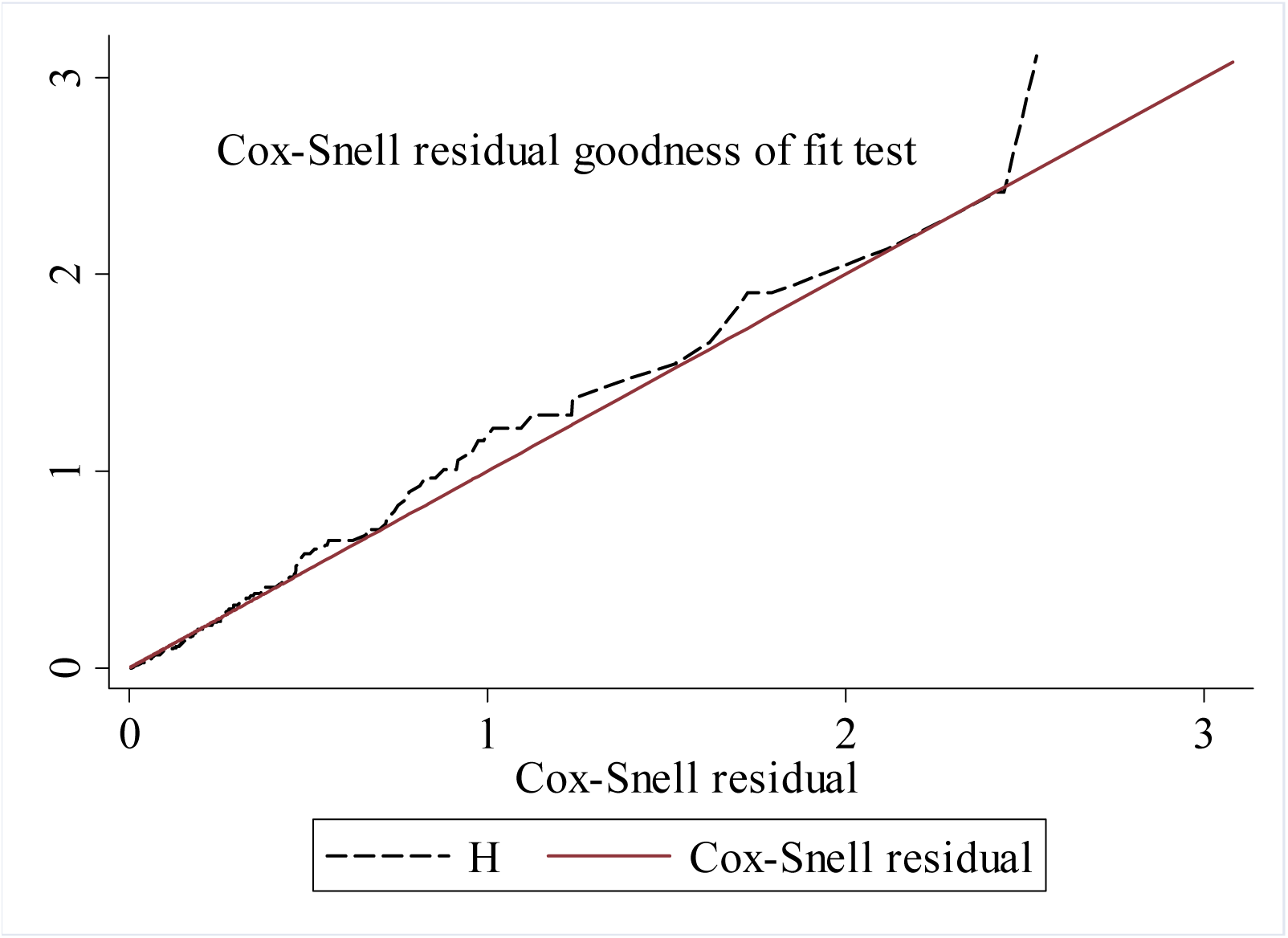
Cox-Snell residual Nelson-Alen cumulative hazard graph on neonates admitted in NICU at Woldia comprehensive specialized hospitals from 1^st^ January 2018 to 22 December 2022.

## Discussion

The objective of this study was to assess the survival time of neonatal death (time to death) and determinants of mortality among neonates admitted to the NICU of Woldia Comprehensive Specialized Hospital. A total of 604 neonates admitted to the NICU were included and reviewed during the study period. Among the admitted neonates, 312 (51.66%) were female, indicating the predominant presence of female neonates in the study population. This finding is consistent with similar studies conducted in Nekemte Specialized Hospital in Western Ethiopia (54.8%) (20), Southern Ethiopia (51.4%) (36) Nigeria (50.9%) (37).

Our study revealed that out of the total 604 neonates included in the study, 117 neonates died during the 3,416 days of follow-up. The median follow-up was 5 days with a minimum of 1 day and the maximum of 28 days. The overall incidence of neonatal mortality was 34.3 per 1000 neonates-days resulting in an overall neonatal mortality rate of 194 (95% CI: 162-225) per 1000 live births. This finding indicates a significant number of newborns admitted to the NICU experienced mortality. The results of our analysis show some similarity to previous studies conducted in different regions of Ethiopia, such as northwestern Ethiopia (151 per 1000 live births), Gondar University Comprehensive Specialized Hospital (173 per 1000 live births), Somali Regional State Ethiopia (186 deaths per 1000 live births), and Wollega (183.4 per 1000 live births) (10,23,38,39). However, our findings are higher than the mortality rates reported in studies conducted at Felege Hiwot referral hospital, Bahir Dar (25) and Jimma University medical center (33).

In comparison to studies conducted in central Ethiopia, where the mortality rate was reported as 233 per 1000 live births (38), and at Mizan Tepi University Teaching Hospital, where it was 227.8 per 1000 live births (15), our study observed a lower proportion of neonatal mortality. Similar findings have been reported in other developing countries, such as India with a rate of 522/1000, Nigeria with 270/1000, and Pakistan with 248/1000 (40).

The variations in these mortality rates may be attributed to several factors, including differences in sample size, study duration, socio-demographic characteristics of the study participants, study settings, healthcare facilities, and methodologies employed. It is important to note that our study specifically focused on neonates admitted to the NICU, which requires high-quality care and often experiences a higher incidence of neonatal mortality. This targeted approach may contribute to the observed differences in mortality rates compared to studies conducted at the community level. Due to the fact that, in referral hospitals different complicated mother can be referred from different parts of the town and neighboring districts and zones suffocated to high flow of complicated cases leads to higher rate of neonatal deaths.

The study revealed a steep increase in the probability of neonatal death during the first seven consecutive follow-up days, as indicated by the Kaplan-Meier estimate curve (Fig 2). The cumulative probability of failure at the end 7^th^, 14^th^ and 28^th^ follow-up days) were 0.17, 0.42 and 0.52 respectively. A significant proportion (56.41%) of deaths occurred within the initial seven days of the neonatal period. These findings are consistent with a similar investigation, which also reported a high occurrence of death during the first week of life. Furthermore, a prior study conducted in Ethiopia (50) found that the highest incidence rate of death occurred in the first week, with a rate of 11.28 per 1000 neonate days of observation (20).

These results align with a study conducted in the NICU of a tertiary hospital in Addis Ababa (22), where approximately 70% of neonatal deaths were registered during the early neonatal period. Similarly, an investigation conducted in a referral hospital in Cameroon reported that over 74.2% of deaths occurred during the early neonatal period (9). These consistent findings suggest that neonates in critical care units face a higher risk of mortality compared to neonates in general. Additionally, there may be a lack of early detection of infections and avoidable neonatal deaths to blame for this.

The results of the multivariable Cox proportional hazard model indicated that ANC checkup during indexed pregnancy was a significant determinant factor for the time to death. Neonates born from mothers who did not receive ANC checkups during pregnancy had twice AHR (95% CI) 1.68(1.12-2.52) increased the hazard of death compared to those who had regular ANC follow-ups. This finding is consistent with a study conducted in Ethiopia (41), which concluded that an increase in ANC visits significantly reduced the risk of neonatal death. Similarly, a study in Kenya highlighted that inadequate or no ANC visits were associated with neonatal mortality (42). The utilization of ANC services is crucial for detecting and treating various conditions, including pregnancy-induced diabetes and hypertension, as well as infections such as syphilis and malaria.

The results of the study revealed that newborns with neonatal sepsis had a 1.87-fold higher mortality risk than neonates without these health issues. Similarly, the hazard of neonatal mortality was 1.64 times higher in the group of neonates with prenatal asphyxia than in the group of neonates without it.

Likewise, a study conducted in in Addis Ababa (31), which consistently reported a higher likelihood of mortality among neonates with sepsis compared to those without. The presence asphyxia, and respiratory distress syndrome (RDS) were significantly associated with neonatal mortality (43). This might be when a newborn has neonatal sepsis, it cannot suckle and exhibits symptoms such as vomiting, diarrhea, poor perfusion, and hypothermia that are linked to a higher chance of dying. The findings of this study emphasize the critical importance of timely detection and management of these health problems to reduce neonatal mortality rates.

## Conclusion and recommendations

A total of 604 neonates admitted to the Neonatal Intensive Care Unit (NICU) during the study years which followed for a total of 3,416 neonatal days from admission to discharge/death. It was found that 14 days was the median time to death. The overall neonatal mortality rate was high, with more than half of the deaths occurring within the first seven days of life. Various factors, ANC checkup, bad obstetrics history of the mother, and specific diagnoses, were found to influence neonatal mortality. Exhaustive efforts should be made to reduce the neonatal mortality. Promoting and giving focused ante natal care (ANC) for every pregnant mother to identify and intervene risks early. Since the study was identified based on the documented data and could not display all factors that were not documented in the patient’s files.

## Data Availability

The data will be available based on the situation

## Acknowledgment

I am very glad to acknowledge the NICU and record office staff of Woldia Comprehensive Specialized Hospital (WCSH) for providing valuable supports during data collection. I also would like to thank data collectors and supervisors.

